# Precision Evidence Bench: Assessing PICO alignment and faithful reporting in clinical AI

**DOI:** 10.64898/2026.10.07.26364976

**Authors:** Arjun Mukerji, Neil Sanghavi, Thomas Z. Lauritzen, Jananee Muralidharan, Saurabh Gombar

## Abstract

Clinicians routinely make decisions for patients whose comorbidities, prior therapies, laboratory abnormalities, or demographic characteristics are not reflected in the supporting evidence, even though these factors can influence treatment selection. Given this gap in the evidence base, we sought to evaluate how AI models respond to clinical questions in which patient characteristics or the treatment comparison and outcome of interest may affect the applicability of available evidence. We developed and publicly released Precision Evidence Bench, a benchmark of 209 synthetic clinical questions with specified population, intervention, comparator, and outcome (PICO) elements. Each answer is assessed using a five-item rubric that evaluates whether a single cited source matches the PICO and whether the answer faithfully reports its sources. The item verdicts determine one of five ratings, ranging from Precision match to Unsupported claim, without requiring a reference answer. We evaluated four frontier large language models (LLMs) using web search across all 209 questions, then repeated the evaluation with additional access to a corpus of precision evidence-based findings (pEBFs) designed to address these evidence gaps, using two corpus sizes. For every model, Precision match rates increased with the amount of precision evidence available, from 13 to 18% with web search alone to 17 to 28% with 100 million pEBFs and 41 to 56% with 500 million pEBFs. These findings suggest that access to matching evidence is an important determinant of LLM performance on precision medicine questions.

## Introduction

Evidence-based treatment decisions require clinicians to assess how published findings apply to the patient in front of them. This can be difficult when patients differ from study participants in their comorbidities, clinical measurements, prior therapies, or other characteristics relevant to treatment selection. Average treatment effects may obscure differences in benefit and harm across patients (Kent et al., 2018), creating a need for evidence that more closely reflects the clinical population and comparison of interest (Gombar et al., 2019). Although trials provide some subgroup evidence, the comparisons and outcomes reported remain limited: among 130 cardiovascular trials published in three major journals, 89 reported at least one subgroup analysis, and only 31 of those reported a subgroup analysis for a secondary outcome (Brand et al., 2021). Inadequate safety reporting across seven medical areas further illustrates the difficulty of obtaining detailed evidence about treatment harms, even before considering patient subgroups (Ioannidis and Lau, 2001). Observational real-world evidence (RWE) can help address some of these gaps, and on-demand consultation services have demonstrated how analyses of previously treated patients can inform individual care and institutional practice (Gombar et al., 2019; Callahan et al., 2021). Patient interest in personalized evidence provides an additional motivation for making such findings accessible (Kaufman et al., 2016; Xu and Wu, 2023).

LLMs equipped with search tools offer a means of retrieving and summarizing this evidence, but the presence of citations does not establish that an answer addresses the clinical question. A response may accurately describe a study involving a different population, comparator, outcome, or follow-up period, leaving the requested comparison unresolved. Alternatively, it may combine findings from several sources into a claim about a comparison that no individual study evaluated. Further errors arise when a model interprets an absence of evidence as evidence of no difference (Altman and Bland, 1995) or makes statements that its cited sources do not support. In an evaluation of medical citation quality, Wu et al. (2025) found that 50–90% of LLM responses were not fully supported by their references and that approximately 30% of statements from GPT-4o with web search were unsupported. Together, these failure modes indicate that clinical evidence retrieval should be evaluated for both relevance to the question and fidelity to the source.

Existing evaluations address aspects of this problem but do not, to our knowledge, assess them together in an open benchmark. Medical question-answering benchmarks primarily evaluate medical knowledge or overall response quality (Jin et al., 2019; Jin et al., 2021; Singhal et al., 2023; Arora et al., 2025), while evaluations of medical retrieval-augmented systems do not require a single source to match all four PICO elements of the question (Xiong et al., 2024; Zakka et al., 2024; Lozano et al., 2024). Citation-quality evaluations assess whether a source supports a statement, which does not necessarily establish its applicability to the patient or treatment comparison being considered (Gao et al., 2023; Wu et al., 2025). Benchmarks for unanswerable questions assess abstention (Rajpurkar et al., 2018; Wen et al., 2025), but the unanswerable items in Med-HALT are nonsensical questions rather than plausible clinical questions for which an answer is not known (Pal et al., 2023). A benchmark focused on patient-specific evidence therefore needs to distinguish accurate reporting of related evidence from evidence that addresses the requested PICO, including when matching evidence may be unavailable.

To address this need, we developed Precision Evidence Bench, comprising 209 synthetic clinical questions and a rubric that evaluates single-source PICO matching and faithful reporting. Because responses are assessed against their own citations and supporting quotations, the rubric does not require a reference answer and can be applied to questions with no known answer. We also report a reference evaluation of four LLMs under three evidence-access conditions, examining whether performance differs according to the availability and location of matching evidence.

## Methods

### Benchmark details

#### Task

Our benchmark focuses on the retrieval of patient-specific evidence and the faithful reporting of that evidence. We implemented it as a single question-answering task. This format is common in clinical LLM evaluation, e.g. MedQA and PubMedQA scenarios in MedHELM (Jin et al., 2021; Jin et al., 2019; Bedi et al., 2026). Questions are posed exactly as written. The model under test is instructed to answer the question and include citations to its sources including verbatim quotes that support its answer.

#### Question generation

A clinical question can be framed by its population, intervention, comparator, and outcome (PICO). We generated questions with known PICO elements so that we could determine whether the retrieved evidence answered the fully specified question that was asked as opposed to only elements of the question. Target outcomes included major clinical events considered primary outcomes in most studies as well as secondary or safety outcomes, such as health care use, laboratory or vital-sign thresholds, and adverse events.

We distinguish general questions from precision questions. A general question asks about the broad population, comparator, and outcome that a trial would study; for example, whether dapagliflozin or sitagliptin is better for adults with type 2 diabetes on metformin. A precision question narrows one or more of these elements below the level that trials report. A precision version of the same example would ask which of the two drugs carries more risk of acute kidney injury among those patients with a history of heart failure. The narrowing can restrict the population by a comorbidity, a clinical measurement, age, or sex, or it can specify a comparator, outcome, or follow-up window.

Precision questions pose a verification problem: many concern comparisons that trials and other literature do not report. We therefore organized questions according to the known availability of matching evidence, including findings from a corpus designed to address precision questions. For 11 questions, a published trial reports the target comparison and outcome in its enrolled population (answer in literature). These serve as positive controls because a system with web search should be able to retrieve the relevant evidence. For 110 questions, the target is a finding in one of our RWE studies (answer in corpus). Of these, 97 target a finding in the RWE study index used in this evaluation (see Reference Evaluation); the other 13 target findings that are not in this index. The 97 questions that targeted an indexed finding were kept only if all four evaluated models reached a full PICO match when the target finding was supplied with the question. For the remaining 88 questions, we know of no corpus finding or trial report (no known answer). These questions assess how a system responds when matching evidence is not known to be available, including whether it substitutes evidence about a different population or comparison.

Candidate questions went through multiple rounds of review to remove those that were clinically implausible, had ambiguous outcomes, or were otherwise confusing, misleading, or ill-formed. In a final round, two physicians reviewed the remaining candidates and commented on their phrasing; applying these comments yielded the final set of 209 questions.

Table 1 describes the composition of the 209 questions. Of these, 53 restrict the population by a comorbidity or clinical measurement and 24 by age or sex alone. The other 132 ask about the whole treated population, and 121 of these instead narrow the comparator, outcome, or follow-up window below the level that trials report. Major clinical events, such as death or heart failure hospitalization, are the target outcome of 32 questions; the remaining 177 ask about health care use (77), a laboratory or vital-sign threshold (26), or another clinical event (74).

**Table 1.** Composition of the benchmark. Patient subgroup: questions restricted by a comorbidity or clinical measurement (53) or by age or sex alone (24), rather than the whole treated population. The target outcome rows count questions by outcome type and sum to the total number of questions. Major clinical event: death, major adverse cardiac events, myocardial infarction, stroke, heart failure hospitalization, or fracture. Health care use: visits, other hospitalizations, length of stay, procedures, and treatment changes. Lab or vital-sign threshold: a named cut-off, such as potassium over 5.1. Other clinical event: adverse events, symptoms, and disease progression. Median question length: 163 characters (range 96–467).

| Characteristic | Questions (N) |
| --- | --- |
| All questions | 209 |
| Patient subgroup | 77 |
| **Target outcome** |  |
| Major clinical event | 32 |
| Health care use | 77 |
| Lab or vital-sign threshold | 26 |
| Other clinical event | 74 |

#### Rubric and Rating

We consider the grading rubric the central contribution of this benchmark. Independently of question selection, it provides a framework for assessing whether a clinical answer rests on matching evidence and reports that evidence faithfully. The criteria can be applied to answers written by clinicians or generated by LLMs.

The rubric scores each answer on five binary items. The first item asks whether a single cited source matches the full PICO of the question, where a single source is one document and a composite of several documents does not qualify. The intervention, comparator, and outcome must match strictly. A population mismatch is acceptable in two cases, when the source population is a superset of the question population or when the answer discloses the mismatch, but a missing required subgroup is not acceptable. Three further items ask whether a single source matches three of the four elements (PIC, PIO, and PCO), each omitting one of the intervention, comparator, or outcome. The fifth item asks whether the answer is faithful to its sources, judged by a materiality test: an error counts against the answer if correcting it would change the conclusion or what a reader believes the evidence found.

Because grading draws only on the question, the answer, and the citations and quotes the model returned, no reference answers are needed. Each item takes the majority verdict of the graders, and the five item verdicts together determine a rating on a five-level scale (Table 2). An answer that is not faithful is rated Unsupported claim (red), regardless of its PICO matches. A faithful answer receives the highest level it meets: Precision match (dark green) when one source matches the full PICO; Partial precision (light green) when two or three of the partial matches hold; Limited precision (yellow) when one partial match holds; and No precision evidence (orange) when none holds.

**Table 2.** From rubric items to ratings. Each item verdict is the majority of the three jurors. The rows are mutually exclusive: every combination of verdicts maps to one rating. “Any” means the item does not affect the rating.

| Rating | Faithful to sources | Full PICO match (one source) | Partial matches (PIC, PIO, PCO) |
| --- | --- | --- | --- |
| Precision match (dark green) | Yes | Yes | Any |
| Partial precision (light green) | Yes | No | 2 or 3 |
| Limited precision (yellow) | Yes | No | 1 |
| No precision evidence (orange) | Yes | No | 0 |
| Unsupported claim (red) | No | Any | Any |

A Precision match rating therefore establishes two properties of an answer: a cited source matches the full PICO of the question, including any required subgroup, and the answer does not misrepresent its sources, as judged against the quotes the model reported. It is equally important to be clear about what the rating does not establish. It does not establish that the reported estimate is correct, that the cited evidence is of high quality or supports a causal conclusion, that the answer is appropriate in clinical practice or offers a sound recommendation for the patient, or that the answer cites a source for each of its claims.

#### Release

We released the benchmark on the Hugging Face Hub as atroposhealth/precision-evidence-bench under a CC BY-SA 4.0 license. This paper evaluates revision v1.0, dated 2026-09-17; the dataset became public on 2026-09-18. The release has four files. The file questions.csv holds the 209 questions, with identifiers prwe-001 to prwe-209. The file rubric.json holds the five rubric items and their grader prompts. The file MANIFEST.json records the row count, a SHA-256 hash for each file, and the source commit. The fourth file is the dataset card. The release does not include the reference labels (such as the answer location of each question), the model results in this paper, or the proprietary corpus.

The dataset card defines a protocol for outside groups. A group poses each question to its system without changes to the wording and asks for an answer with citations. A grader scores each answer on the five rubric items, with rubric.json as its instructions. The card does not prescribe a grader. When there are multiple graders, a majority vote is used to decide each item. The group reports results per item or as the five-level rating specified above.

#### Reference Evaluation

We evaluated four models, each with its vendor’s web search tool: GPT-5.6 Sol and GPT-6 Astra (OpenAI API), Claude Opus 5, and Gemini 3.8 Flash. Each model was presented with every question under three conditions. In the “Without Alexandria” condition, the model received the question and web search only; the other two conditions added the Alexandria retrieval tool at two corpus sizes.

Alexandria is Atropos Health’s proprietary library of real-world evidence, generated with the RWE study methodology described in detail by Coyle et al. (2026). When this study was run in mid-September 2026, Alexandria held 500 million precision evidence-based findings (pEBFs). The retrieval tool searches four sources in parallel: the RWE study index, PubMed, a Prognostogram library (Callahan et al., 2021), and ASCO guidelines. The Alexandria, 500M pEBFs condition used the full RWE study index, whereas in the Alexandria, 100M pEBFs condition a deterministic hash of the study identifier restricted the index to one fifth of its studies.

No model was trained or fine-tuned on Alexandria content; the models reached Alexandria only through the retrieval tool, which they called while answering. All three conditions are therefore forms of retrieval-augmented generation (Lewis et al., 2020) in which the model calls search tools as it answers (Yao et al., 2023), and for a given model they differ only in the tools offered, plus an instruction to use the Alexandria tool when it is available.

A jury of three LLMs graded each answer: Claude Opus 5, GPT-5.6 Sol, and Gemini 3.7 Flash. Each juror saw the question, the answer, and the quotes the model reported, and scored the answer on all five items of the v1.0 rubric in a single call, with the rubric text unchanged. Each item took the majority verdict of the three jurors, and the item verdicts set the rating as described in Rubric and rating. We ran each model under each condition once, in September 2026. During rubric development, clinicians reviewed verdicts on 20 responses, and one clinician agreed with the jury majority on 16.

For each model and condition, we report the share of answers at each rating level across all 209 questions (Figure 1) and within each answer location (Table 3).

**Table 3.** Precision match and Unsupported claim by answer location. Percent of answered questions for each model and condition. Gemini 3.8 Flash answered 97, 110, and 110 of the corpus questions; 10 of the 11 literature questions in each condition; and 84, 87, and 88 of the no-known-answer questions.

| Answer location | Model | Precision match, without | Precision match, 100M | Precision match, 500M | Unsupported claim, without | Unsupported claim, 100M | Unsupported claim, 500M |
| --- | --- | --- | --- | --- | --- | --- | --- |
| Answer in corpus (n=110) | GPT-5.6 Sol | 16.4 | 26.4 | 83.6 | 10.9 | 8.2 | 10.9 |
|  | GPT-6 Astra | 10 | 20 | 62.7 | 2.7 | 2.7 | 3.6 |
|  | Claude Opus 5 | 10.9 | 30.9 | 72.7 | 10.9 | 16.4 | 16.4 |
|  | Gemini 3.8 Flash | 11.3 | 30.9 | 81.8 | 63.9 | 30 | 10 |
| Answer in literature (n=11) | GPT-5.6 Sol | 72.7 | 72.7 | 72.7 | 18.2 | 9.1 | 9.1 |
|  | GPT-6 Astra | 54.5 | 54.5 | 63.6 | 0 | 0 | 0 |
|  | Claude Opus 5 | 72.7 | 63.6 | 54.5 | 0 | 9.1 | 9.1 |
|  | Gemini 3.8 Flash | 60 | 80 | 70 | 40 | 20 | 30 |
| No known answer (n=88) | GPT-5.6 Sol | 12.5 | 14.8 | 18.2 | 11.4 | 10.2 | 11.4 |
|  | GPT-6 Astra | 11.4 | 8 | 10.2 | 13.6 | 6.8 | 9.1 |
|  | Claude Opus 5 | 15.9 | 15.9 | 11.4 | 6.8 | 6.8 | 14.8 |
|  | Gemini 3.8 Flash | 13.1 | 17.2 | 14.8 | 40.5 | 42.5 | 35.2 |

**Figure 1.**
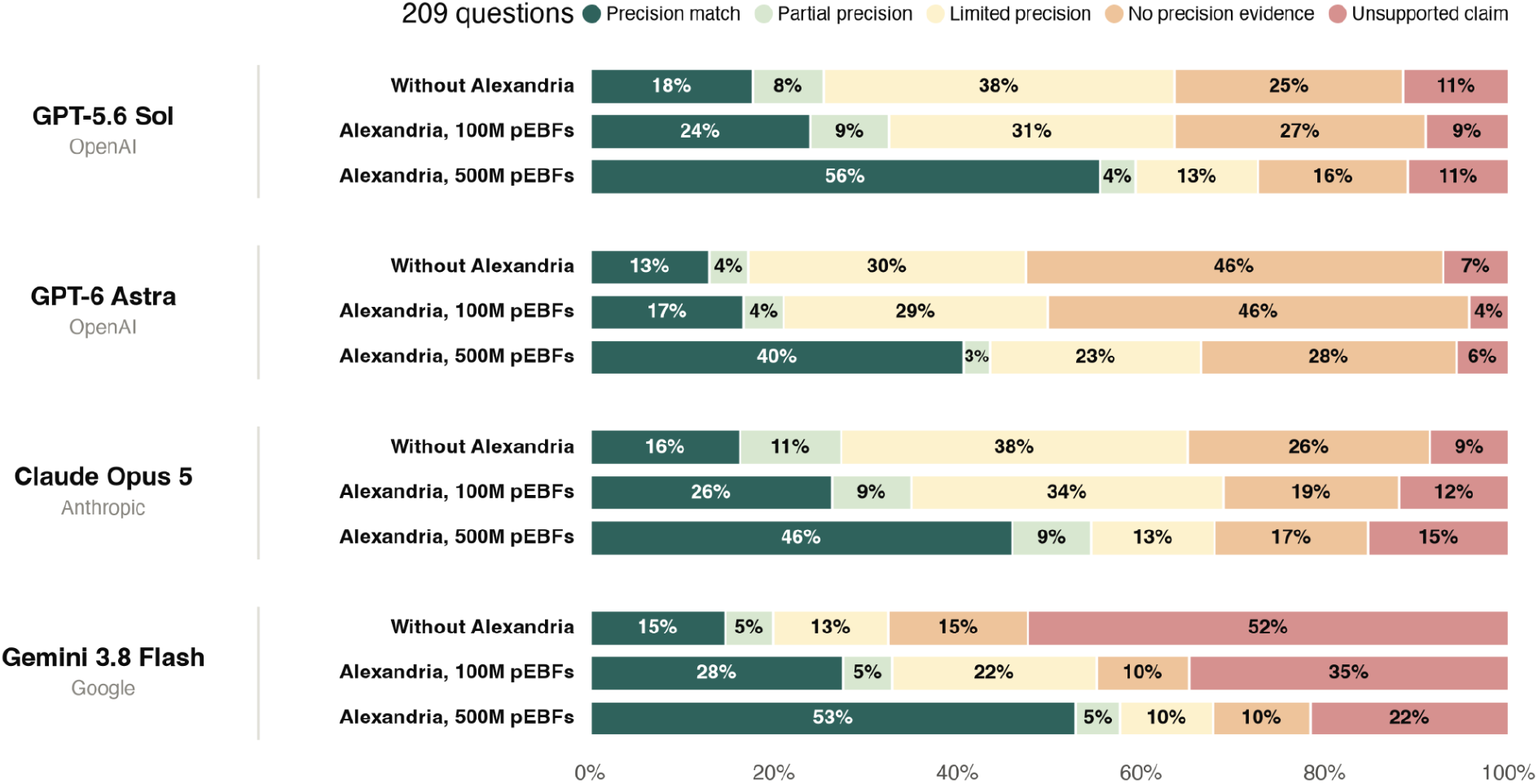
More precision evidence, better answers. Distribution of ratings for four LLMs under three conditions, pooled over the 209 questions. Without Alexandria: the model’s vendor web search tool. Alexandria, 100M pEBFs: web search and the Alexandria retrieval tool, with the RWE study index restricted to about 20% of its studies. Alexandria, 500M pEBFs: web search and the Alexandria retrieval tool over the full library. pEBF, precision evidence-based finding. Ratings combine the majority verdicts of a three-LLM jury (Claude Opus 5, GPT-5.6 Sol, Gemini 3.7 Flash) on the five items of the v1.0 rubric. Model labels name the models as served through each vendor’s API. Shares are over answered questions; Gemini 3.8 Flash answered 191, 207, and 208 of the 209 questions in the three conditions. Labels are rounded by the largest-remainder method so that each bar sums to 100%; bar widths use unrounded shares.

## Results

Figure 1 shows the distribution of ratings for each model under each condition over the 209 questions. Gemini 3.8 Flash returned no answer to 18, 2, and 1 questions in the three conditions; we report its shares over the questions it answered.

Access to more precision evidence raised Precision matches for every model. Without Alexandria, the Precision match rate ranged from 13% (GPT-6 Astra) to 18% (GPT-5.6 Sol). It ranged from 17% to 28% at 100M pEBFs and from 41% to 56% at 500M pEBFs, a 2.8- to 3.6-fold increase over web search alone. For every model, the second step was larger than the first: +20 to +32 percentage points from 100M to 500M pEBFs, against +4 to +13 points from web search alone to 100M.

The 100M subset kept the target study for 21 of the 97 questions whose target is an indexed finding. At 100M pEBFs, Precision match reached 62–90% on these 21 questions and 3–13% on the 76 questions whose target study was dropped, in line with the 8–13% those 76 scored without Alexandria. At 500M pEBFs, with every target present, the two groups reached 67–86% and 64–91%.

Without Alexandria, most answers from three models were faithful but either answered a nearby question (limited precision, yellow) or did not find relevant evidence (no precision evidence, orange). Limited precision and No precision evidence held 63% of answers from GPT-5.6 Sol, 76% from GPT-6 Astra, and 64% from Claude Opus 5. These answers reported their sources without material error, but the sources differed from the question in population, comparator, or outcome. At 500M pEBFs, the share of these two levels fell by 33, 25, and 34 percentage points, against Precision match gains of 38, 28, and 30 points. Unsupported claims stayed between 4% and 11% for GPT-5.6 Sol and GPT-6 Astra across conditions and rose from 9% to 15% for Claude Opus 5. For these three models, evidence access changed whether an answer matched the question more than whether it was faithful to its sources.

Gemini 3.8 Flash failed in a different way. Without Alexandria, 52% of its answers were rated as unsupported and 28% were faithful but rated Limited precision or No precision evidence. The share of answers rated as unsupported fell to 35% at 100M pEBFs and to 22% at 500M pEBFs, while the Precision match share rose from 15% to 53%. For this model, more evidence came with fewer unfaithful answers as well as more matches.

As expected, the pooled gain in Figure 1 came from the questions whose answer is in the corpus. Of the questions that reached Precision match at 500M pEBFs but not without Alexandria, 90–97% per model had their answer in the corpus. For these 110 questions, Precision match ranged from 10% to 16% without Alexandria, from 20% to 31% at 100M pEBFs, and from 63% to 84% at 500M pEBFs, a gain of +53 to +70 percentage points over web search alone. The fall in Unsupported claim for Gemini 3.8 Flash came from these questions: 64% without Alexandria, 30% at 100M pEBFs, and 10% at 500M pEBFs. On questions with no known answer, its Unsupported claim share stayed between 35% and 43%.

For the 11 questions with the answer in the trial literature, Alexandria added little. Precision match without Alexandria ranged from 55% to 73%, and Alexandria did not raise it. Changes across conditions amounted to two questions or fewer per model, in both directions.

Among the 88 questions with no known answer, Precision match rates remained between 8% and 18% across conditions. Changes were no greater than six percentage points, with no consistent direction across models. These matches came disproportionately from the 30 no-known-answer questions about the whole treated population. Those questions reached Precision match in 13% to 33% of answers, against 2% to 12% for the 58 questions restricted to a subgroup. The rubric’s allowances for a population mismatch (a superset population, or a mismatch the answer discloses) offer a route to a match on these 30 questions; on the other 58, a missing required subgroup is not allowed.

## Discussion

Precision Evidence Bench evaluates whether clinical AI responses cite evidence that addresses the requested comparison and faithfully report what that evidence shows. The reference evaluation demonstrates why both dimensions are needed: three of the four models frequently provided faithful summaries of evidence that did not answer the requested PICO. Expanded retrieval access was associated with higher Precision match rates across all four models but did not consistently improve faithfulness. Within this evaluation, changes associated with evidence access were larger than differences among models under the same condition. Analyses stratified by answer location indicate that improvements were concentrated among questions with retrievable target findings, supporting the importance of evidence availability while also reflecting the benchmark’s construction and selection procedures.

The distinction between source faithfulness and clinical relevance was particularly apparent for three of the four models. Their most common responses without Alexandria accurately represented the cited evidence but did not address the requested population, comparator, or outcome. Citation-support assessments may accept such responses because the source supports the statement being made (Rashkin et al., 2023; Gao et al., 2023; Wu et al., 2025), even though the statement leaves the clinical question unresolved. Evaluating PICO matching alongside faithfulness makes this limitation explicit. The partial-match items can also help identify which PICO element is missing, although the present analysis reports the combined response ratings.

The two dimensions also responded differently to expanded evidence access. For GPT-5.6 Sol, GPT-6 Astra, and Claude Opus 5, gains in matching were accompanied by limited or unfavorable changes in Unsupported claim rates, indicating that retrieval of more relevant evidence does not necessarily improve how faithfully a model reports it. Gemini 3.8 Flash showed improvements in both dimensions, with the reduction in unsupported responses concentrated among corpus questions. Assessing matching and faithfulness separately therefore provides a more informative account of system performance than either measure alone: a system may retrieve a relevant study and misrepresent it, or accurately summarize a study that does not answer the question.

Several features of question construction limit the interpretation and generalizability of these results. All questions were synthetic, and 84 of 209 (40%) used templated wording that named the study comparison, outcome, and follow-up window. Moreover, the 97 indexed corpus questions were selected only when all four evaluated models achieved a full PICO match after receiving the target finding. Because this screen used the same models and jury as the evaluation, performance when the target was retrievable partly reflects the selection process. The no-known-answer label was also heuristic. A question received it when our corpus contained no study of the comparison and the question asked about a secondary or safety outcome within a patient subgroup, a combination that trials rarely report. Only 58 of the 88 questions are restricted to a subgroup; the other 30 ask about the whole treated population, so the rule does not cover them. We did not search the literature exhaustively for these questions, so the label should not be taken as proof that matching evidence does not exist. Finally, the literature group included only 11 questions, and the public release omits answer-location labels.

The evaluation also depends on the judgments of an LLM jury. Using jurors from different model families may reduce dependence on a single judge (Verga et al., 2024), but changing the panel could change the ratings. Two jurors were also among the evaluated models, creating the possibility of self-preference described in previous work (Zheng et al., 2023; Panickssery et al., 2024); we observed no indication of this behavior, and GPT-5.6 Sol was the strictest judge of its own responses.

Each model was evaluated once per condition, and vendor-served models may change over time. Public release of the questions also creates the possibility that future models will have encountered them during training (Sainz et al., 2023).

For clinicians seeking evidence about a specific patient population and treatment comparison, an accurate summary of related literature may still leave the decision unsupported. Precision Evidence Bench provides an open framework for evaluating whether clinical AI responses cite evidence that satisfies the requested PICO and report it faithfully. In this reference evaluation, access to matching evidence was associated with substantial improvements in benchmark performance. Further evaluation using clinician-authored questions and independent assessment of the cited sources would help establish how these findings translate to routine clinical use.

## Data Availability

All data produced are available online at https://huggingface.co/datasets/atroposhealth/precision-evidence-bench/tree/main

## Acknowledgements

Use of large language models. Large language models (Claude Opus 5.5, Anthropic, accessed via the Claude Code interface) were used to assist in drafting and editing the text of this manuscript, including composing and revising prose from author-supplied results, and copyediting for clarity and concision. All authors carefully reviewed, verified, and approved every passage of AI-assisted text and take full responsibility for the accuracy, integrity, and originality of the submitted work.

All authors are employed by Atropos Health, which owns Alexandria and its retrieval tool and developed the question generators and rubric.

